# Development and validation of an enzyme immunoassay for detection and quantification of SARS-CoV-2 salivary IgA and IgG

**DOI:** 10.1101/2021.09.03.21263078

**Authors:** Veronica P. Costantini, Kenny Nguyen, Zoe Lyski, Shannon Novosad, Ana C. Bardossy, Amanda K. Lyons, Paige Gable, Preeta K. Kutty, Joseph D. Lutgring, Amanda Brunton, Natalie Thornburg, Allison C. Brown, L. Clifford McDonald, William Messer, Jan Vinjé

## Abstract

Oral fluids offer a non-invasive sampling method for the detection of antibodies. Quantification of IgA and IgG antibodies in saliva allows studies of the mucosal and systemic immune response after natural infection or vaccination. We developed and validated an enzyme immunoassay (EIA) to detect and quantify salivary IgA and IgG antibodies against the prefusion-stabilized form of the SARS-CoV-2 spike protein. Normalization against total antibody isotype was performed to account for specimen differences, such as collection time and sample volume. Saliva samples collected from 187 SARS-CoV-2 confirmed cases enrolled in 2 cohorts and 373 pre-pandemic saliva samples were tested. The sensitivity of both EIAs was high (IgA: 95.5%; IgG: 89.7%) without compromising specificity (IgA: 99%; IgG: 97%). No cross reactivity with seasonal coronaviruses was observed. The limit of detection for SARS-CoV-2 salivary IgA and IgG assays were 1.98 ng/mL and 0.30 ng/mL, respectively. Salivary IgA and IgG antibodies were detected earlier in patients with mild COVID-19 symptoms than in severe cases. However, severe cases showed higher salivary antibody titers than those with a mild infection. Salivary IgA titers quickly decreased after 6 weeks in mild cases but remained detectable until at least week 10 in severe cases. Salivary IgG titers remained high for all patients, regardless of disease severity. In conclusion, EIAs for both IgA and IgG had high specificity and sensitivity for the confirmation of current or recent SARS-CoV-2 infections and evaluation of the IgA and IgG immune response.

## INTRODUCTION

Severe acute respiratory syndrome coronavirus 2 (SARS-CoV-2), the causative agent of the coronavirus disease 2019 (COVID-19) pandemic, is a Betacoronavirus related to Severe Acute Respiratory Syndrome Coronavirus (SARS-CoV) and Middle East Respiratory Syndrome Coronavirus (MERS-CoV) [1-3]. As of July 30, 2021, SARS-CoV-2 infections has caused more than 200 million cases worldwide, and an estimated 4.2 million deaths. The clinical spectrum of SARS-CoV-2 infection ranges from asymptomatic infection to symptomatic disease [4]. The high proportion of asymptomatic individuals not only results in a high transmission rate, but also suggests differences in the host immune response compared to other coronaviruses [5]. Since the duration of immunity to SARS-CoV-2 dictates the overall course of the pandemic as well as post-pandemic strategies, a comprehensive understanding of the relationship between systemic and mucosal antibody responses becomes important. Because the oral and nasal cavities are considered main sites for SARS-CoV-2 entry and replication, locally produced mucosal antibodies may protect against infection. Therefore, saliva samples can be used as a non-invasive tool for virus detection as well as measuring the immune response (mucosal and systemic) [6].

Salivary antibody levels can be 100 to 1000-fold lower than serum levels [7]. Salivary IgG is mainly derived from serum by leakage across capillaries and enters saliva through gingival crevices. At mucosal membranes, IgA is the main immunoglobulin class and is found most often in the secretory form (sIgA). Within 2-3 weeks after onset of disease, SARS-CoV-2–specific IgG antibodies can be detected in saliva, persist for at least 9 months, and show high correlation with serum antibody levels in most COVID-19 patients [8-11]. Salivary IgA antibodies, on the contrary, rapidly increase 1 week after onset of disease, become undetectable 4-5 weeks later, and show a moderate correlation with serum levels [8, 9]. Saliva provides a non-invasive collection method, easy to implement in remote areas and community settings without a need for extensive training. These features, while additionally evaluating both mucosal and systemic immune responses, make salivary antibody testing an ideal approach to evaluate population immunity, transmission, asymptomatic infections, and vaccine performance. We previously demonstrated the value of saliva-based antibody assays to evaluate immune responses mounted against norovirus [12]. In this manuscript, we describe the development and validation of an enzyme immunoassay (EIA) to quantitatively evaluate the presence of SARS-CoV-2–specific IgA and IgG antibodies in saliva and describe the salivary immune response to SARS-CoV-2 mounted in different cohorts of infected patients.

## MATERIALS AND METHODS

### Saliva samples

A total of 333 saliva samples were collected from 187 participants who had tested positive for SARS-CoV-2 by real-time reverse transcription polymerase chain reaction (rRT-PCR) [13] or antigen test in two cohorts **(Figure 1)**. In cohort I, 235 samples were collected from a) 113 participants at a single time point and b) 32 participants on a weekly basis for 4-5 weeks (n=122) after diagnosis. In cohort II, 98 saliva samples were collected from 42 participants with either asymptomatic (n=8), mild (n=29) or severe disease (n=5) at different times after the onset of disease (range 0-203 days). Disease severity was defined according to the World Health Organization criteria [4] and clinical data were obtained using a standardized questionnaire. In addition, 373 pre-pandemic archived samples collected between 2009-2010 were included as negative controls [12].

**Figure 1:**
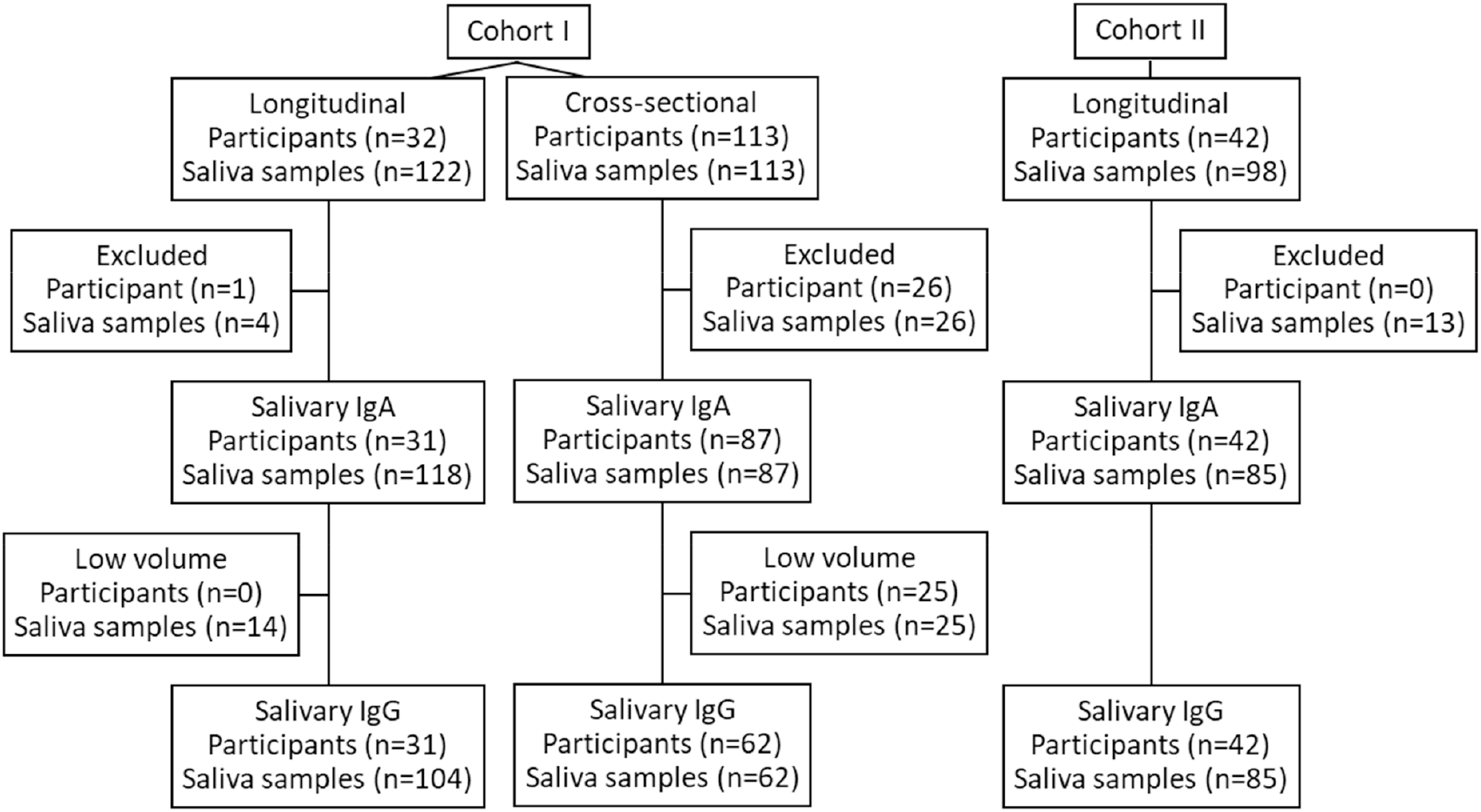
Specimen collection and testing. A total of 333 saliva samples were collected from 187 participants who had tested positive for SARS-CoV-2 by rRT-PCR or antigen test. In cohort I, 113 participants provided a single sample and 32 participants provided samples on a weekly basis for 4-5 weeks after diagnosis. In cohort II, 42 participants provided saliva samples at different times after onset of diseases.

### Specimen collection and processing

Saliva was collected at least 30 minutes after consumption of food or liquids. Pre-pandemic archived saliva samples were collected using the Oracol saliva collection device, processed, and stored at -80°C according to the manufacturer’s instructions (Malvern Medical Developments, Worcester, UK). Cohort I samples were collected using the Oracol S14 collection device by gently rubbing the swab along the gumline around the entire mouth for approximately 1 minute. This collection device specifically harvests gingival crevicular fluid, which resembles serum composition [14]. Saliva samples collected by the Oracol swabs were separated by centrifugation (10 min at 1500 x g), transferred to the attached microtube (10 μl -200 μl), and stored at -80°C until analysis. For cohort II, participants were asked to cough deeply and spit into a collection cup containing virus isolation media (PBS plus 2% FBS, Gentamicin, Amphotericin B). Saliva samples were clarified by centrifugation (10 min at 3000 x g), aliquoted and stored at -80°C until analysis. All samples collected during the pandemic were inactivated by gamma radiation (2 × 10^6^ rads) prior to testing [15]. Samples were initially tested for SARS-CoV-2–specific salivary IgA. If enough sample volume (≥ 100μl) was available, samples were tested for SARS-CoV-2–specific salivary IgG.

### Enzyme immunoassay (EIA) for detecting SARS-CoV-2-specific IgA and IgG in saliva

Convalescent sera from 3 SARS-CoV-2 patients with IgA and IgG antibodies against SARS-CoV-2 spike protein were used for the initial assay development. The pre-fusion stabilized ectodomain of SARS-CoV-2 spike (S) that was used in the assays was obtained from in suspension adapted HEK-293 cells as described previously [16]. Antigen concentrations ranging from 0.125-1.00 μg/ml in PBS and 1:1,000 to 1:20,000 diluted horse radish peroxidase (HRP)-conjugated goat anti-human IgA or IgG were initially tested. Positive and negative controls (SARS-CoV-2 convalescent serum and pre-pandemic saliva samples, respectively), as well as blank controls (only blocking buffer) were also included in each run. All volumes were 100 μl per well, except where indicated. All washes were performed 3 times with 250 μl of PBS/0.05% Tween-20 (PBST) using a BioTek 405 plate washer. All dilutions were prepared in blocking buffer (5% (w/vol) powdered milk/PBST). All incubations were carried out for 1 h at 37°C except where indicated. All concentrations and incubation times were optimized to maximize the optical density (OD) difference between pre-pandemic negative samples and SARS-CoV-2 convalescent sera.

Immunol 2 HB flat 96 well-plates (Fisher Scientific) were coated with 100 µl SARS-CoV-2 spike protein (0.5 µg/ml in PBS, row A to D, positive coated wells) or PBS (row E-H, negative coated wells) and incubated overnight at 4°C in a humidified chamber. Plates were washed, blocked with 200 μl /well blocking buffer for 2 h at 37°C. After blocking, plates were washed three times and 4-fold serial dilutions (1:10-1:160) of each saliva sample were added to both antigen and PBS coated wells. Plates were incubated, washed, and bound antibodies were detected using HRP-conjugated goat anti-human IgA (Sera Care) diluted 1:4,000 in blocking buffer or anti-IgG (Sera Care), diluted 1:16,000. After incubation, plates were washed again before adding 100 ul of 3,3′,5,5′-Tetramethylbenzidine (TMB) substrate (Sera Care). The colorimetric reaction was stopped 5 min later by adding 100 µl Stop solution (Sera Care). The plates were read at 450 nm and 630 nm using an Epoch2 instrument (BioTek). After background correction, ODs were calculated (OD _450nm_ -OD_630nm_) and the adjusted OD value (OD value positive coated well minus OD value negative coated well) was determined.

### Enzyme immunoassay (EIA) for detecting total IgA and IgG in saliva

For the detection of total IgA, Immunol 2 HB flat 96 well-plates (Fisher Scientific) were coated with goat anti-human IgA (α-chain) (0.5 µg/ml in PBS, rows A to F, positive coated wells) or PBS 1x (rows G-H, negative coated wells) and incubated overnight at 4°C in a humidified chamber. Plates were washed and blocked with blocking buffer (200 μl /well) for 2 h at 37°C. Three dilutions (1:1,280, 1:5,120 and 1:20,480) of each saliva sample were prepared. A standard curve for IgA was prepared by serial dilution of purified human IgA (Sigma-Aldrich). After incubation, plates were washed and saliva samples were added to rows A-D. Purified human IgA dilutions were added to both positive coated (anti IgA, rows E-F) and negative coated (PBS, rows G-H) wells. Plates were incubated, washed, and bound antibodies were detected using 1:4,000 diluted HRP-conjugated goat anti-human IgA (Sera Care). After incubation, plates were washed again before adding TMB substrate (Sera Care). The colorimetric reaction was stopped 5 min later by adding Stop solution (Sera Care).

The same plate design and steps were used for IgG. Plates were coated with goat anti-human IgG (γ-chain) at 0.5 µg/ml in PBS. A standard curve for IgG was prepared by serial dilutions of purified human IgG (Sigma-Aldrich) and bound antibodies were detected using 1:16,000 diluted HRP-conjugated goat anti-human IgG (Sera Care).

### Sensitivity and specificity

The sensitivity (ability to identify samples with antibodies to SARS-CoV-2), and the specificity (ability to identify samples without antibodies to SARS-CoV-2) were defined as the values for which there is 95% probability that the estimated value can be obtained [17]. For IgA EIA validation, 373 SARS-CoV-2 rRT-PCR negative saliva samples (pre-pandemic) collected from healthy adults (2009-2010) and 44 saliva samples from SARS-CoV-2 rRT-PCR-confirmed cases were used. Similarly, for validation of the IgG assay, 373 pre-pandemic saliva samples and 68 saliva samples from confirmed cases were used. Saliva samples were collected ranging from 0 to 63 days after diagnosis.

### Limit of detection

The limit of detection (LOD) was defined as the lowest predicted value for which there is 95% probability that an estimated value can be obtained. To determine the LOD for the IgA (or IgG) EIA, 5 IgA (or IgG) positive SARS-CoV-2 saliva samples were 4-fold serially diluted (1:10-1:10,240). The adjusted OD values were extrapolated from the linear portion of the IgA (or IgG) standard curve. The LOD was determined from the lowest concentration of antibody above the cut off value (mean of the adjusted OD values for the pre-pandemic samples + 3SD).

### Analytical specificity

To evaluate potential cross reactivity between antibodies against the spike protein of SARS-CoV-2 and other human coronaviruses, convalescent serum from patients positive for the common human coronaviruses [HuCoV 229 (n=2), HuCoV NL-63 (n=1), HuCoV OC43 (n=7) and HuCoV HUK1 (n=1)] were included.

### Data analysis

Descriptive statistics for continuous variables are presented as median plus interquartile ranges (IQR). For categorical variables, n (%) was used for descriptive statistics and differences were evaluated by Chi-Square or Fisher’s exact test. Non-parametric data from more than 2 groups were compared by Kruskal-Wallis test. Receiver operating characteristics (ROC) analysis was used to evaluate sensitivity and specificity of each assay when establishing a cut off for positivity. Each saliva sample was tested at 2 different dilutions. To quantify total and SARS-CoV-2 specific antibodies, adjusted OD values from serially diluted purified human IgA (or IgG) were plotted against concentration and fit to a sigmoidal 4 parameter logistic model. Total and SARS-CoV-2 -specific salivary IgA (or IgG) titers were extrapolated from the linear portion of the IgA (or IgG) standard curve. To account for participant differences (e.g., severity of illness, immunocompetency, collection time, antibody secretion levels), SARS-CoV-2 specific IgA or IgG were normalized to 100 µg of total IgA or IgG, respectively. When virus-specific antibodies could not be detected, but total antibodies were detected, SARS-CoV-2-specific IgA (or IgG) per 100 µg of total IgA (or IgG) were arbitrarily assigned as half the lower limit of detection, based on the standard curve of purified human IgA [18]. Statistical analysis was performed using GraphPad Prism version 8.0 (GraphPad Software, San Diego, California USA, www.graphpad.com), and *p*-values <0.05 were considered significant.

### Ethics statement and Disclaimer

This activity was reviewed by CDC and was conducted consistent with applicable federal law and CDC policy (See, e.g., 45 C.F.R. part 46.102(l)(2), 21 C.F.R. part 56; 42 U.S.C. §241(d); 5 U.S.C. §552a; 44 U.S.C. §3501 et seq.). The study was approved by the Institutional Review Boards of the Oregon Health & Science University (IRB# 21230) (cohort II). Participants that were cognitively or decisionally impaired were excluded. Written informed consent was obtained from each participant. The findings and conclusions in this article are those of the authors and do not necessarily represent the official position of the Centers for Disease Control and Prevention.

## RESULTS

We developed and validated EIAs for the quantitative detection of SARS-CoV-2-specific salivary IgA and IgG antibodies in saliva. The final assay conditions are summarized in Table 1. Pre-pandemic saliva (negative for SARS-CoV-2 antibodies) and convalescent serum from COVID-19 patients were used for the initial standardization of both EIAs. All reagent concentrations [SARS-CoV-2 spike protein, anti-human IgA (α-chain) or IgG (γ-chain), HRP- labeled secondary antibody, purified human IgA and purified human IgG concentration for standard curves] and incubation times were optimized to reduce background, obtain maximum anti-SARS-CoV-2 specific signal, and to maximize the optical density difference between pre-pandemic negative samples and SARS-CoV-2 convalescent sera.

**Table 1:**
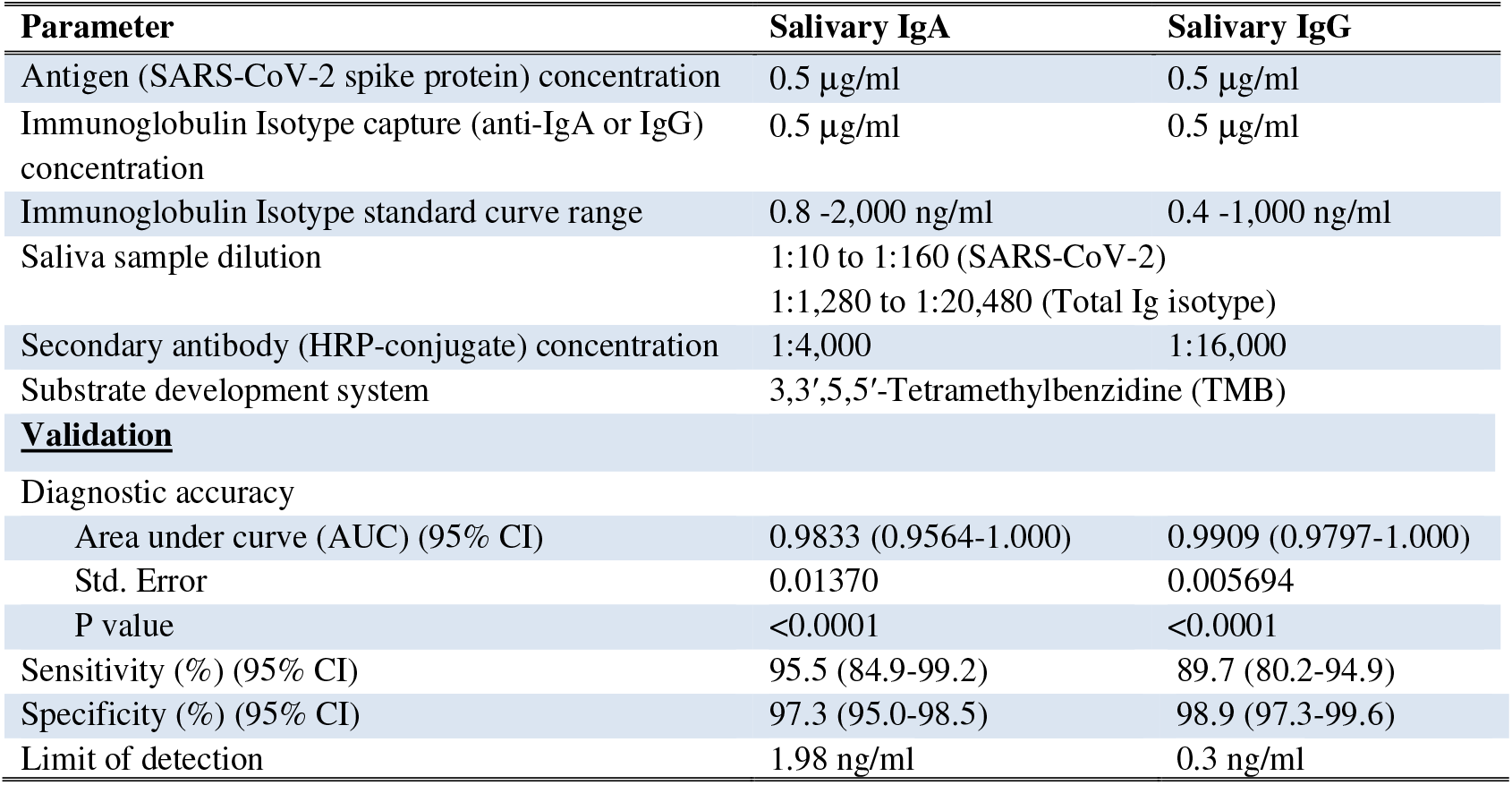
Parameter optimization and assay validation

### Clinical sensitivity and specificity

A dilution of 1:10 of true negative (pre-pandemic) and true positive (SARS-CoV-2 rRT-PCR– confirmed cases) saliva samples resulted in OD values with no background signal in the negative coated wells (PBS coat). Adjusted OD values for both SARS-CoV-2-specific IgA and IgG antibodies from true positive samples were significantly higher in saliva from rRT-PCR confirmed cases than from pre-pandemic samples (**Figure 2A**, p< 0.0001). ROC analysis was applied to determine the cut off value that maximizes sensitivity and specificity (**Figure 2B and C)**. For the IgA EIA, the assay specificity improved from 91.5% (95% CI: 88.2%-93.9%) to 97.3% (95% CI: 95.0%-98.5%), when the cut off value increased from one to three standard deviations (*SD*) above the mean 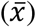 of the pre-pandemic samples, whereas the sensitivity remained at 95.5% (95% CI: 84.9%-99.2%). For the IgG EIA, a significant increase of the specificity from 97.9% (95% CI: 96.6%-99.3%) to 98.9% (95% CI: 97.3%-99.6%) (p<0.05) was achieved when the cut off value was modified from 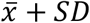 to 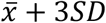. Although the sensitivity slightly decreased from 91.2% (95% CI: 82.1%-95.9%) to 89.7% (95% CI: 80.2%-94.9%), this difference was not significant (p=0.0625). Based on these data we used a cut off value of 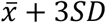 of the adjusted OD values from pre-pandemic saliva samples for both the IgA and IgG EIAs assays. The overall diagnostic accuracy was 98.3% (95% CI:95.6%-100%) and 99.1 (95% CI:98.0%-100%) for the IgA and IgG assays, respectively (Table 1).

**Figure 2:**
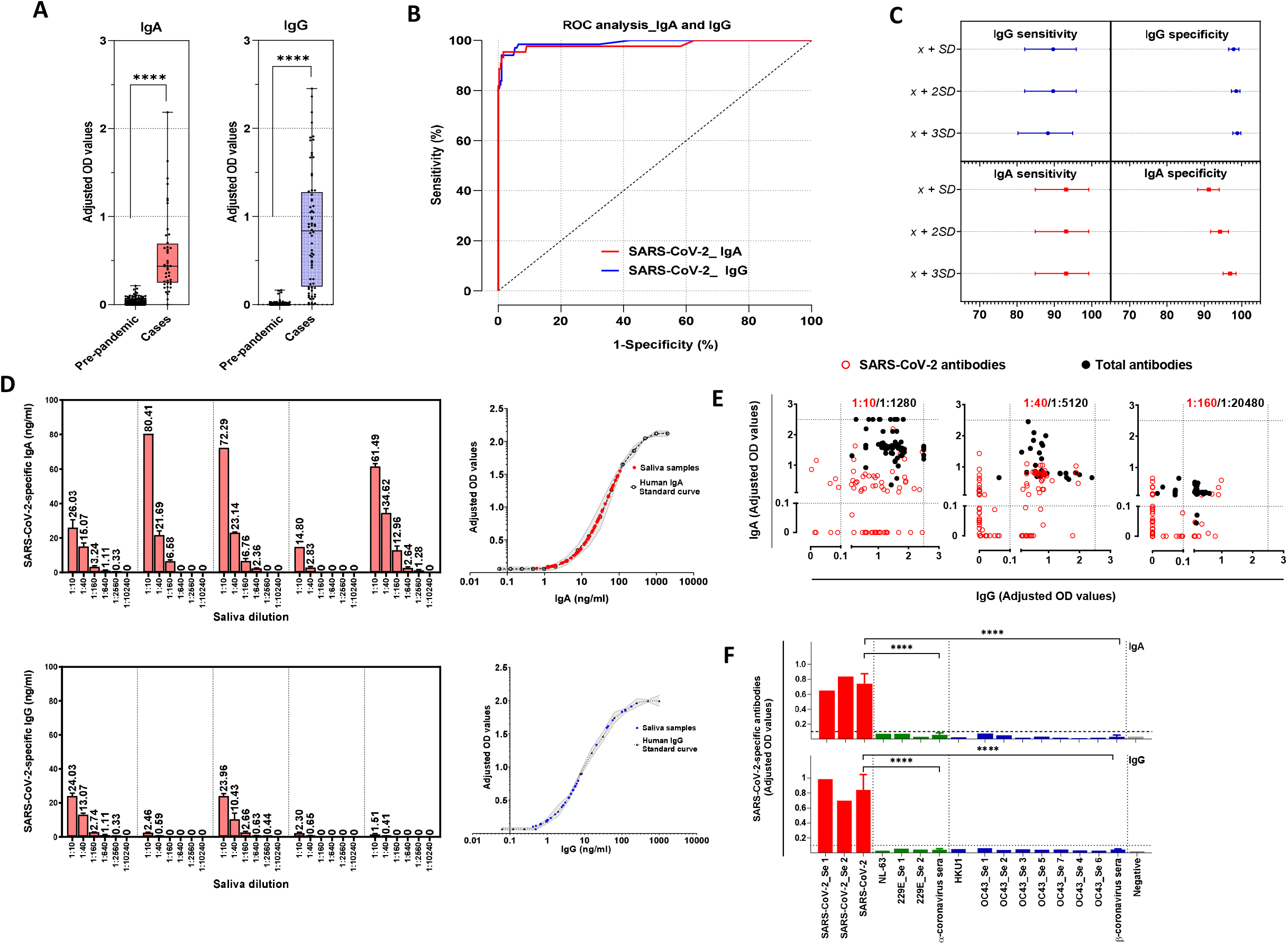
Development and validation of enzyme immunoassays for the detection of salivary IgA and IgG against SARS-CoV-2. Saliva samples were collected from SARS-CoV-2 confirmed cases (IgA, n=44; IgG, n=68). Pre-pandemic samples were collected between 2009-2010 (n=373). Saliva samples were diluted 1:10 and added to a 96-well plate precoated with SARS-CoV-2 S antigen. **(A)** Sample adjusted OD values for SARS-CoV-2 IgA and IgG. **(B)** Receiver operating curves for each assay were constructed with data from SARS-CoV-2 confirmed cases, as well as pre-pandemic samples. The optimal cut off value to differentiate cases from controls was set as the maximum Youden’s index (sensitivity + specificity - 1). **(C)** Sensitivity (95% CI) and specificity (95%CI) for three different cut off values [mean 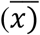 of the pre-pandemic samples plus standard deviations (*SD*)] were considered for each antibody isotype. **(D)** To assess the limit of detection for each isotype, non-linear regression was performed using 5 saliva samples serially diluted in 2 independent experiments. Adjusted OD values were extrapolated from IgA or IgG standard curve for each sample and dilution. SARS-CoV-2 specific antibody concentrations (ng/ml) are indicated on top of each bar. **(E)** Saliva sample dilution evaluation. Saliva samples from SARS-CoV-2 confirmed cases (n=66) were tested for total (dilutions 1:1,280 to 1:20,480, black dots) and SARS-CoV-2 (dilutions 1:10 to 1:160, red dots) antibodies. False negative rates were calculated for each dilution. The dotted line represents the calculated cutoff value (OD 0.1) discriminating between positive and negative samples, and the upper limits of quantification (OD 2.5) above which detectors on the plate reader are saturated. **(F)** Specificity of antigens against a panel of seasonal coronavirus convalescent serum. SARS-CoV-2 S antigen was tested against convalescent serum from confirmed SARS-CoV-2 cases (red), seasonal coronavirus cases prior to pandemic (green and blue) and pre-pandemic negative samples (gray) for IgG and IgA. The horizontal dotted line represents the calculated cutoff value discriminating between positive and negative samples based on ROC analysis in (C).

### Limit of detection

For both IgA and IgG antibody assays, a sigmoidal four-parameter logistic curve was fitted to the resulting adjusted OD values to yield a standard curve of antibody concentration versus OD **(Figure 2D)**. The lower limit of detection for the assays was 1.98 ng/ml for SARS-CoV-2 IgA antibodies and 0.3 ng/ml for SARS-CoV-2 IgG antibodies. At the upper limits of quantification, above which the detectors on the plate reader for total IgA and IgG antibodies are saturated, samples were tested at higher dilutions (< 1:1,280) to fit within the linear portion of the sigmoidal curve, without compromising the detection of antibodies against SARS-CoV-2. We tested 3 dilutions for SARS-CoV-2 and total antibodies. (Table 1).

At 1:10 dilution, 42 saliva samples tested positive for SARS-CoV-2 IgA whereas at 1:40 and 1:160 dilutions, 16/42 (38%) and 25/42 (59%) false negatives were detected. For total IgA antibodies, 10/42 (24%) saliva samples were above the upper limit of quantification at 1:1,280 dilution, whereas at 1:5,024 dilution 42/42 (100%) were detected and 1:20,480 dilution 2/42 (5%) false negative were detected. At 1:10 dilution, 60 saliva samples tested positive for SARS-CoV-2 IgG, whereas at 1:40 and 1:160, 33/60 (55%) and 40/60 (67%) false negatives were detected. For total IgG antibodies, 12/60 (20%) saliva samples were above the upper limit of quantification at 1:1,280 dilution, whereas at 1:5,024 dilution 60/60 (100%) were detected and 1:20,480 dilution 7/60 (12%) false negative were detected **(Figure 2E)**. Based on these data we chose to test each saliva sample at 1:10 and 1:40 dilution for SARS-CoV-2 antibodies and 1:1,280 and 1:5,120 dilution for total antibodies.

### Analytical specificity

To evaluate the potential cross reactivity with antibodies against seasonal coronaviruses (229E, HKU1, OC43, NL63), convalescent serum samples from confirmed seasonal coronavirus patients were tested for the presence of SARS-CoV-2 IgA and IgG **(Figure 2F)**. At the lowest serum dilution (1:100), both SARS-CoV-2 IgA and IgG adjusted OD values were significantly lower in seasonal α-coronavirus (0.032±0.021 and 0.045±0.01), and seasonal β-coronavirus (0.058±0.024 and 0.044±0.014) serum than SARS-CoV-2 serum (0.742±0.133 and 0.841±0.203) (p<0.0001), suggesting that our assays show no cross-reactivity with antibodies against seasonal coronaviruses.

### Kinetic and magnitude of SARS-CoV-2 specific antibody response in saliva after infection

Cohort I yielded 205 saliva samples from 118 SARS-CoV-2 confirmed cases available to be tested for SARS-CoV-2-specific salivary IgA and 166 saliva samples for which enough saliva was available were also tested for IgG **(Figure 1)**. The mean time between SARS -CoV-2 diagnosis to saliva collection was 22.9 days (median 20; IQR: 14-31.75; range: 2–65 days). Overall, 86.4% (102/118) of participants had measurable salivary antibodies for one (IgA: 19.5%; n=23/118; IgG: 20.3%, n=24/118) or both isotypes (46.6%, 55/118). Conversely, 13.6% (16/118) participants tested negative for both SARS-CoV-2 salivary IgA and IgG. During the first week after diagnosis, the positivity rate for both IgA and IgG was 28.6% (2/7 samples). For IgA, positivity rate rapidly increased to 62.1% at week 2, peaked at week 4 (68.6%) and decreased to 0% at week 9 and week 10. At week 3, the positivity rate for IgG was 80%, peaked at 100% at week 8 and remained positive to the end of the study (week 10) **(Figure 3A)**. The median amount of SARS-CoV-2-specific IgA was 0.1 (IQR:0.1-49.44) ng/100 µg of total IgA at week 1 and increased to 7.95 (IQR: 0.1-25.74) ng/100 µg of total IgA at week 3 (IQR: 0.1-25.74). After fluctuating at week 4 and week 6, the IgA levels became undetectable at week 9. The salivary IgG titers increased between week 1 (median: 0.1; IQR:0.1-14.32) and week 5 (median: 123.4; IQR:39.32-269.5), increased to 162.9 (IQR:31.93-322.9) ng/100 µg of total IgG at week 6 and then remained stable throughout the follow-up period. Compared to week 1 significant differences between IgG levels were observed at week 5 (p<0.05) and week 6 (p=0.01) **(Figure 3B)**.

**Figure 3:**
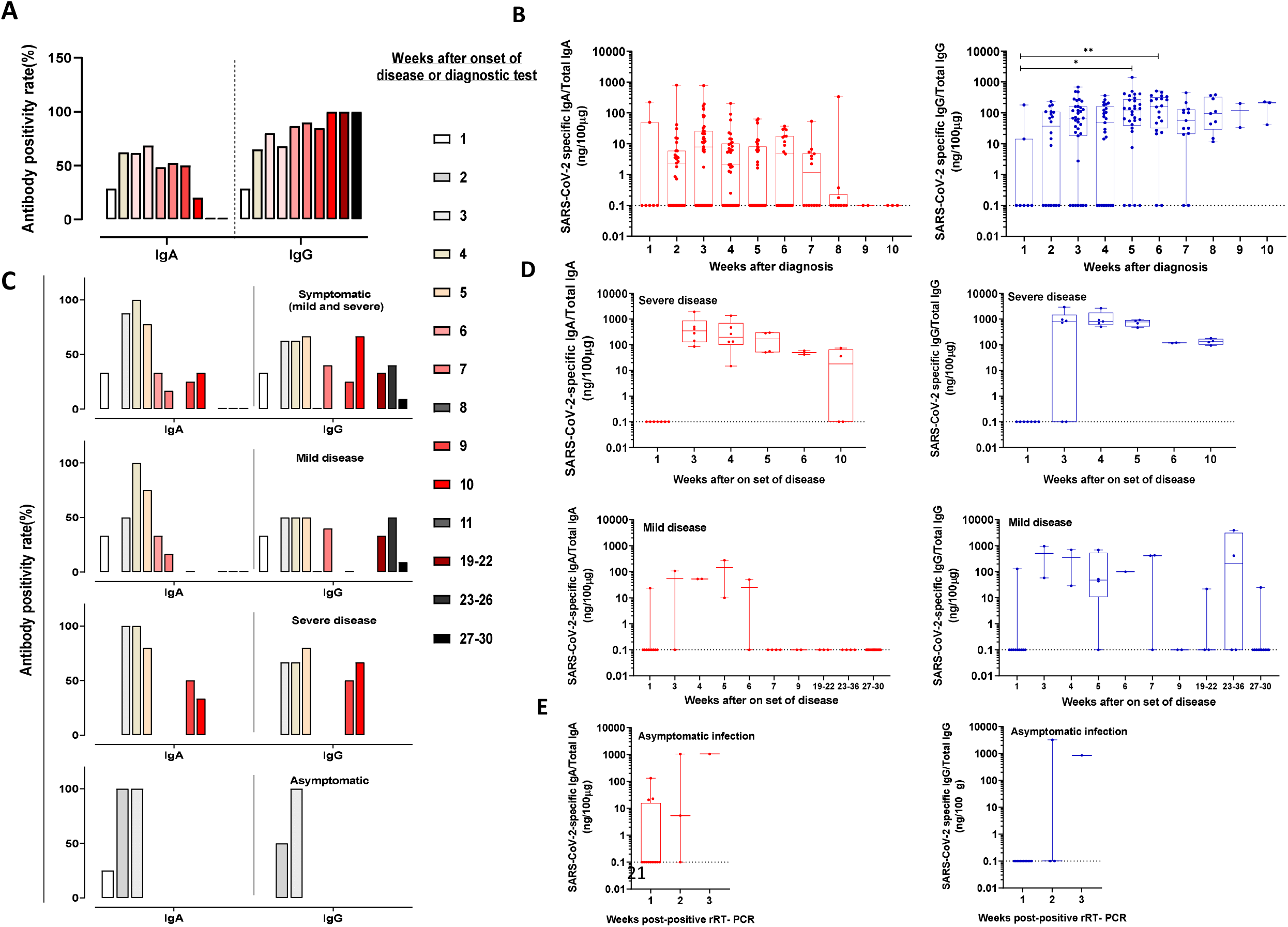
Kinetics and magnitude of SARS-CoV-2 specific salivary antibody response after infection. Positivity rate after **(A)** positive diagnostic test in cohort I (118 and 93 participants for IgA or IgG, respectively) or **(C)** onset of disease in cohort II (42 participants). Empty column indicates no sample available. Positive rate equal 0% is indicated by an underscore. **(B and D)** SARS-CoV-2-specific salivary antibody levels in COVID-19 patients. SARS-CoV-2 salivary IgA and IgG levels after a positive diagnostic test on day 0 (B, Cohort I, 205 and 166 samples for IgA and IgG, respectively) or on set of disease (D, Cohort II, 85 saliva samples for IgA and IgG). SARS-CoV-2 antibodies were normalized to 100 μg of total salivary IgA or IgG, respectively to account for differences among participants, collection time and secretion levels. When SARS-CoV-2 specific salivary antibodies were not detected, but total antibodies (IgA or IgG, respectively) were present, normalized SARS-CoV-2 salivary antibody levels were arbitrarily assigned to 0.1 for visualization purpose (dotted line)

In the cohort II, 85 saliva samples from 42 rRT-PCR-positive participants who were either asymptomatic (n=8), had mild (n= 29) or severe (n= 5) clinical COVID-19 disease symptoms were tested for IgA and IgG antibodies **(Table 2 and Figure 1)**. In the group of patients with mild symptoms the mean time between onset of disease to sample collection was 146.8 days (median: 178; IQR: 81.25-200.8; range 1-208 days) and 25.6 days (median: 18; IQR: 14.5-40.5; range:12-60 days) in the group with severe symptoms.

**Table 2:** Demographics and clinical characteristics of the SARS-CoV-2 patients from cohort II

| Characteristic | Asymptomatic<br>(n=8) | Mild disease <sup>a</sup><br>(n=29) | Severe disease <sup>ab</sup><br>(n=5) | Total <sup>b</sup><br>(n=42) |
| --- | --- | --- | --- | --- |
| Age (y), median (IQR)<sup>c</sup> | 44.5 (25.2-49.7) | 62.0 (44.0.-65.5)* | 63(49.0-74.0)* | 56.0(43.5-65) |
| Sex(male/female) | 2/6 | 10/19 | 2/3 | 14/28 |
| **SARS-CoV-2 infection severity, n,**<br>**(%)<sup>d</sup>** |  |  |  |  |
| **<u>Ambulatory</u>** |  |  |  |  |
| No limitation of activities | 8(100) | - | - | 8 (19.0) |
| Limitation of activities | - | 22 (75.9) | - | 22(52.4) |
| **<u>Hospitalized</u>** |  |  |  |  |
| No oxygen therapy | - | 1(3.4) | - | 1 (2.4) |
| Oxygen therapy <sup>e</sup> | - | 6(20.7) | - | 6 (14.3) |
| Non-invasive ventilation | - | - | 3(60.0) | 3 (7.1) |
| Intubation (mechanical ventilation) | - | - | - | - |
| Ventilation +additional organ<br>support. | - | - | 2(40.0) | 2(4.8) |
| **Level of care at saliva sampling,**<br>**n, (%)** |  |  |  |  |
| Outpatient | 8(100) | 22(75.9) | - | 30(71.4) |
| Hospitalized | - | 7(24.1) | 5(100) | 12 (28.6)** |
| **Symptoms** |  |  |  |  |
| Fever >=100.4 F | - | 16(55.2) | 1(20.0) | 17(40.5) |
| Chills | - | 9(31.0) | 1(20.0) | 10 (23.8) |
| Weakness | - | 15(51.7) | 3(60.0) | 18 (42.9) |
| Muscle aches | - | 12(41.4) | 1(20.0) | 13 (31.0) |
| Runny nose | - | 8(27.6) | 1(20.0) | 9 (21.4) |
| Sore throat | - | 9(31.0) | - | 9 (21.4) |
| Cough | - | 23(79.3) | 3(60.0) | 26 (61.9) |
| Shortness of breath | - | 21(72.4) | 5(100.0) | 26 (61.9) |
| Nausea | - | 5(17.2) | - | 5 (11.9) |
| Vomiting | - | - | - | - |
| Headache | - | 11(37.9) | 1(20.0) | 12 (28.6) |
| Diarrhea | - | 5(17.2) | 2(40.0) | 7 (16.7) |
| Abdominal Pain | - | 4(13.8) | - | 4 (9.5) |
| Rash | - | - | - | - |
| Other | - | 19(65.6) | 1(20.0) | 20 (47.6) |

<sup>a</sup> Disease severity (mild vs severe) was defined according to the World Health Organization Classification [4]
<sup>b</sup> Categorical values were compared using Fisher exact test or Chi-square for more than 2 groups. Continues variable were compared by Kruskal-Wallis test. \* p<0.05; \*\* p<0.001
<sup>c</sup> Interquartile range
<sup>d</sup> Clinical spectrum of SARS-CoV-2 infection according to World Health Organization Classification [4]
<sup>e</sup> Oxygen by mask or nasal prong

Overall, for cohort II, the positivity rate for IgA showed a clear pattern with increasing values during week 1-3, a peak at week 4, whereas the positivity rate for IgG was less defined. **(Figure 3C)**. For patients with mild disease symptoms, the positivity rate for IgA and IgG was 33.3% at week 1 after onset of symptoms. The IgA titer rapidly increased and peaked at week 4 (100%), sharply decreased to 0% at week 9 and remained negative until week 30. The positivity rate for IgG peaked at 50% at week 3, fluctuated for several weeks and returned to 0% 30 weeks after onset of disease. In patients with severe clinical symptoms, the positivity rate for IgA and IgG were consistently higher than in mild cases at each time point. The positivity rate for IgA peaked at week 3 (100%) then slowly decreased to baseline at week 10 whereas IgG peaked (80%) at week 5 and remained elevated until the end of the study (week 10).

In cohort II, salivary IgA levels in patients with severe clinical symptoms peaked at week 3 (median:346; IQR: 128.2-855.9) and remained positive for the entire study (10 weeks), whereas salivary IgG titers peaked at week 4 (median: 810.5; IQR: 598.2-1778) and remained positive until week 10. In patients with mild disease, salivary IgA titers peaked later, at week 5 (median: 143.6; IQR: 9.94-277.3) and became undetectable at week 7, while salivary IgG antibodies peaked earlier at week 3 (median: 507.9; IQR: 58-957). Although with fluctuating values, salivary IgG titers remained detectable until week 36 after onset of disease. In asymptomatic participants (n=8), salivary IgA titers were detected 1 week earlier than IgG. Overall, during the first 6 weeks after onset of symptoms, salivary IgA and IgG titers were higher in patients with severe symptoms compared to patients with mild symptoms **(Figure 3D)**.

## DISCUSSION

We report the development and validation of in-house EIAs to quantitatively assess the presence of SARS-CoV-2-specific IgA and IgG in saliva and showed its value to describe the salivary immune response after natural SARS-CoV-2 infection. Both the IgA and IgG assays were highly accurate, sensitive, and specific. The sensitivity of both assays was high (IgA: 95.5%; IgG: 89.7%) without compromising specificity (IgA: 99%; IgG: 97%). Other published studies have shown high sensitivity for IgG (88-98.4%), but low (17-59%) for IgA, with a high specificity for both isotypes (96-100%) [8-11]. Difference in sensitivity between our and other IgA assays could be explained by the assay platform, type of sample, or collection time after infection. The limit of detection of 1.98 ng/ml for SARS-CoV-2 IgA and 0.3 ng/ml for SARS-CoV-2-specific IgG suggest that both assays are suitable for the detection of salivary antibodies in samples collected early after infection up to several weeks after recovery.

Overall, we detected SARS-CoV-2-specific IgA and IgG responses as early as 1 week after onset of disease or diagnosis when disease data were not available. The IgA positive rate decreased to zero after 10 weeks, whereas IgG positivity rate remained high for at least up to 30 weeks. Similarly, data from previous cross-sectional studies showed detectable SARS-CoV-2 -specific IgA and IgG levels in saliva 2–4 weeks after onset of symptoms, with only IgG response antibodies persisting beyond 60 days [8, 9]. A different study reporting results from single saliva samples collected <3 to 9 months after onset of disease showed a consistently high IgG positivity, but a significant decrease of IgA [10]. In a longitudinal study including 95 participants, the mean time from disease onset to IgG detection in saliva was 9-11 days and IgG antibodies remained detectable until day 90 [19].

In cohort II, when no saliva sample was collected during the first week after onset of disease, we assumed that participants were negative for both SARS-CoV-2-specific IgA and IgG. Salivary IgA levels peaked 3 weeks after onset of disease and remained elevated until at least week 10 in participants with severe disease symptoms. Conversely, in patients with mild disease salivary IgA levels were transient, peaked late at week 5 and returned to baseline levels at week 7. Participants with severe disease showed a later (week 4) peak for IgG than those with mild disease, and both remained positive until the end of the study. These initial increases in salivary IgA and IgG were similar in asymptomatic individuals, although the number of samples available after 1 week was limited. Our data agree with the study by Pisanic *et al*., where salivary IgA and IgG antibodies reached a peak at 3 weeks post infection, but only IgG remained above baseline levels for at least 60 days after onset of symptoms [9]. Another group found similar IgG and IgA kinetics in serum from mild and severe cases [20]. Several studies reported higher correlation for IgG than IgA when testing paired serum and saliva samples [8, 9], leading to question the advantage of testing IgA in saliva. Early SARS-CoV-2 humoral immune responses are dominated by IgA antibodies which remained detectable in saliva for a longer time than in serum (days 49 to 73 post-symptoms). IgA antibodies also contribute to virus neutralization to a greater extent compared to IgG, although they circulate for shorter time than IgG [21]. Given the less invasive collection method, and the transient presence of IgA in contrast to long-lasting IgG, testing for IgA in saliva might allow for a better understanding of the timing of infection.

Our study has several limitations. First, data on the onset and severity of disease was not always available. Second, collection of saliva samples did not always start on day 0 after onset of disease, not all participants provided samples at the same time points, and a limited number of longitudinal samples from asymptomatic individuals were available. Third, due to low sample volume, IgG testing could not be performed on all saliva samples. Fourth, saliva samples were not screened for the presence of antibodies against SARS-CoV-1, MERS-CoV or seasonal coronaviruses. We did test convalescent sera positive for seasonal coronavirus and showed no cross-reactivity. Others have shown some cross reactivity with SARS-CoV-1 and MERS-CoV which will be relevant only if these viruses circulate in the same population tested with our assay for antibodies against SARS-CoV-2. Finally, paired serum samples to correlate the immune response were not available; therefore, correlation between saliva and serum could not be evaluated.

In summary, we developed and validated EIAs that are sensitive and specific to detect SARS-CoV-2 specific IgA and IgG antibodies in saliva. Significant fluctuations of salivary IgA and IgG antibodies levels were observed after infection. Detection of salivary antibodies may serve as an easy-to-employ screening method for population and transmission studies as well as evaluation of vaccine response, especially when collection of blood is challenging or not feasible.

## Data Availability

Datasets generated and analyzed during the current study are available from the corresponding author on reasonable request.

## ACKNOWLEDGMENT

We are grateful to the participants for their willingness to participate during the challenging time of the pandemic. We would like to thank Jennifer Harcourt and Mohammed Rasheed at the Respiratory Immunology Team and Owen Herzegh and David Petway at the Division of Scientific Resources for their support with the gamma irradiation process. We would like to thank John Jernigan, Sujan Reddy, Farrell Tobolowsky, Kelly Hatfield, and Signature HealthCARE for their assistance with recruiting participants. We are also grateful to Peter Sullivan, Matthew Strnad, Felicity Coulter, and Sarah Siegel for logistical and administrative support for patient consent and sample processing at OHSU.

